# New Epidemiological Model Suggestions Revealing Size of Epidemics Based on the COVID-19 Pandemic Example: Wavelength Models

**DOI:** 10.1101/2020.04.07.20056432

**Authors:** Tevfik Bulut

## Abstract

The main purpose of the study is to introduce the wavelength models developed to measure the size of outbreaks based on the COVID-19 example. In this way, the wavelengths of the outbreaks can be calculated, ensuring that the outbreaks are valid, reliable and easy to follow at the national and international level. Wavelength models consist of approved case, death, recovered case and net wavelength models. Thus, the size of the outbreak can be measured both individually and as a whole. COVID-19 cases of 181 countries were used to demonstrate the application of the models. The prominent findings in the applied wavelength models are as follows: the countries with the highest case wavelength are USA, Italy, Spain and Germany, respectively. However, Italy ranks first in the death wavelength, followed by Spain, the USA and France. On the other hand, China has taken the first place in the recovered case wavelength. This country was followed by Spain and Germany and Italy, respectively. Based on all these wavelength models mentioned, net wavelength lengths are calculated. According to the findings of net wavelengths obtained, Canada ranked first, followed by United Kingdom, USA and Italy, respectively.

## Introduction

In the coronavirus pandemic, which has deeply influenced the world, the total number of confirmed cases has exceeded one million and the number of those who died has exceeded fifty-two thousand as of April 2, 2020 [4]. While this is the case, the world has been involved in an intensive study from preventive measures to therapeutic measures to combat the coronavirus outbreak [8, 9,10, 11, 15].

Coronaviruses (CoV) are a large family of viruses that cause many diseases, from the common cold to more serious diseases such as the Middle East Respiratory Syndrome (MERS-CoV) and Severe Acute Respiratory Syndrome (SARS-CoV) [14]. This new virus is called coronavirus because its surface protrusions are crowned, and this family of viruses are single chain, positive polarity, enveloped RNA viruses. Coronavirus disease (COVID-19) was discovered in Wuhan, China’s Hubei Province in December 2019. This disease is transmitted from person to person through breathing [6, 10, 12, 13].

Within the scope of this study, a mathematical model was developed to measure the extent of outbreaks in other outbreaks, especially coronavirus pandemics, which deeply affected countries. With this model, it was aimed to calculate the wavelength of the outbreaks and to facilitate the follow-up of the outbreaks on a reliable and valid basis at the national and international level. Data sets retrieved from “The Humanitarian Data Exchange (HDX)” website were used to demonstrate the implementation of the models [4].

## Methodology

In this study, the size of the outbreaks, especially the coronavirus outbreak, was determined with the mathematical models developed in the current data set, and the size of the outbreaks was measurable, as well as the opportunity to compare outputs of these modes within the country and between countries.

To calculate wavelengths, data sets was taken from the Human Data Exchange (HDX) platform which is the one of the websites of OCHA (United Nations Office for the Coordination of Humanitarian Affairs [4]. The number of cases in the data sets covers the period from 2020-01-22 until 2020-04-02 (including this date). Data sets having the extension csv (comma-seperated value) were combined because they consisted of 3 different datasets, including confirmed cases and deaths, and recovered coronavirus cases. The number of cases in the data sets follows a cumulative course. Microsoft Excel 2016 and R Programming language was used in the analysis [2,3].

Since there are duplicates in the time series for countries in the data sets, a unique time series was created within each country using data mining techniques. Descriptive statistics of the World coronavirus (COVID-19) cases in the data set are given in Table 1.

**Table 1:**
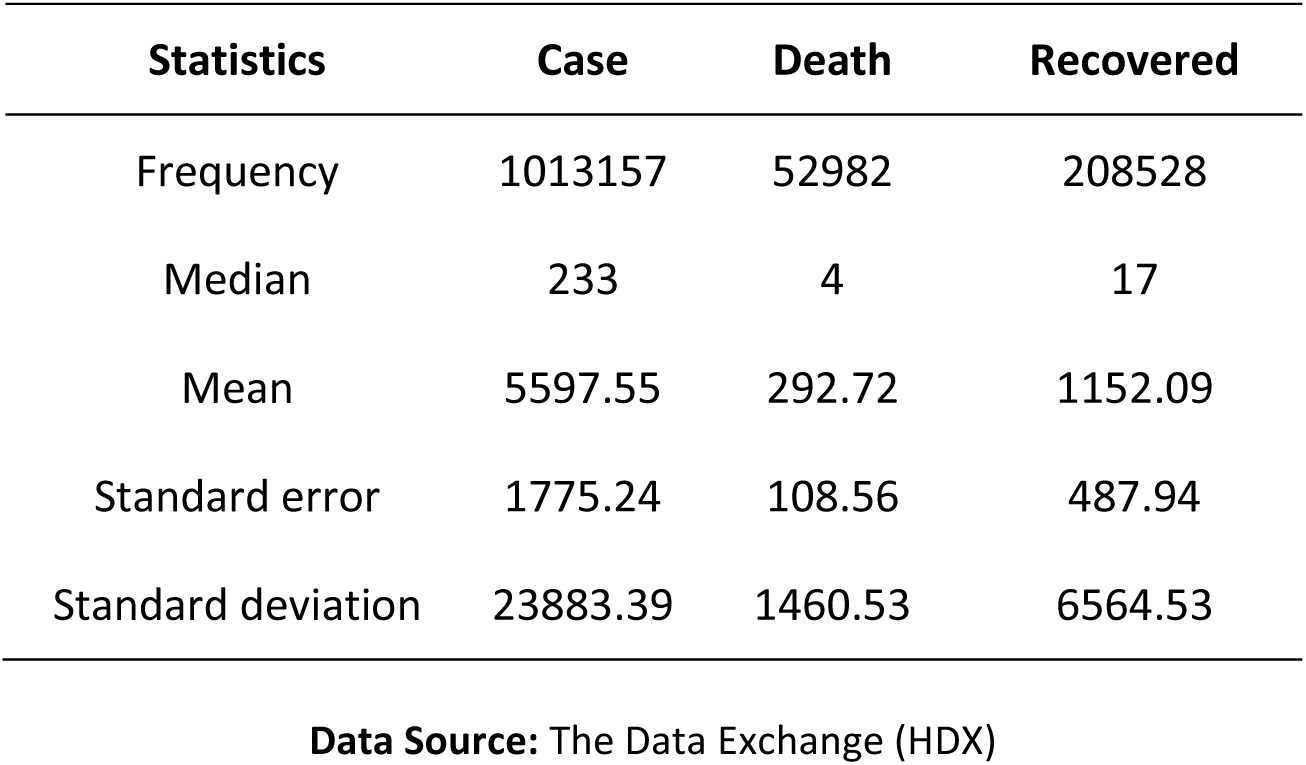
Descriptive Statistics

| Statistics | Case | Death | Recovered |
| --- | --- | --- | --- |
| Frequency | 1013157 | 52982 | 208528 |
| Median | 233 | 4 | 17 |
| Mean | 5597.55 | 292.72 | 1152.09 |
| Standard error | 1775.24 | 108.56 | 487.94 |
| Standard deviation | 23883.39 | 1460.53 | 6564.53 |

**Data Source:** The Data Exchange (HDX)

The course of the COVID-19 cases in the world has been presented in graphs from 2020-01-22 until 2020-04-02 (including this date), which is handled separately on the basis of approved cases, deaths and recovered cases. The blue dashed line parallel to the x axis in the graphs shows the average of the cases. First, the course followed by the approved cases in Figure 1 is given. There was a sharp increase in the number of cases approved according to Figure 1 at the end of the first 60-day period. After this time period, it was observed that the number of approved cases was above the average number of cases.

**Figure 1:**
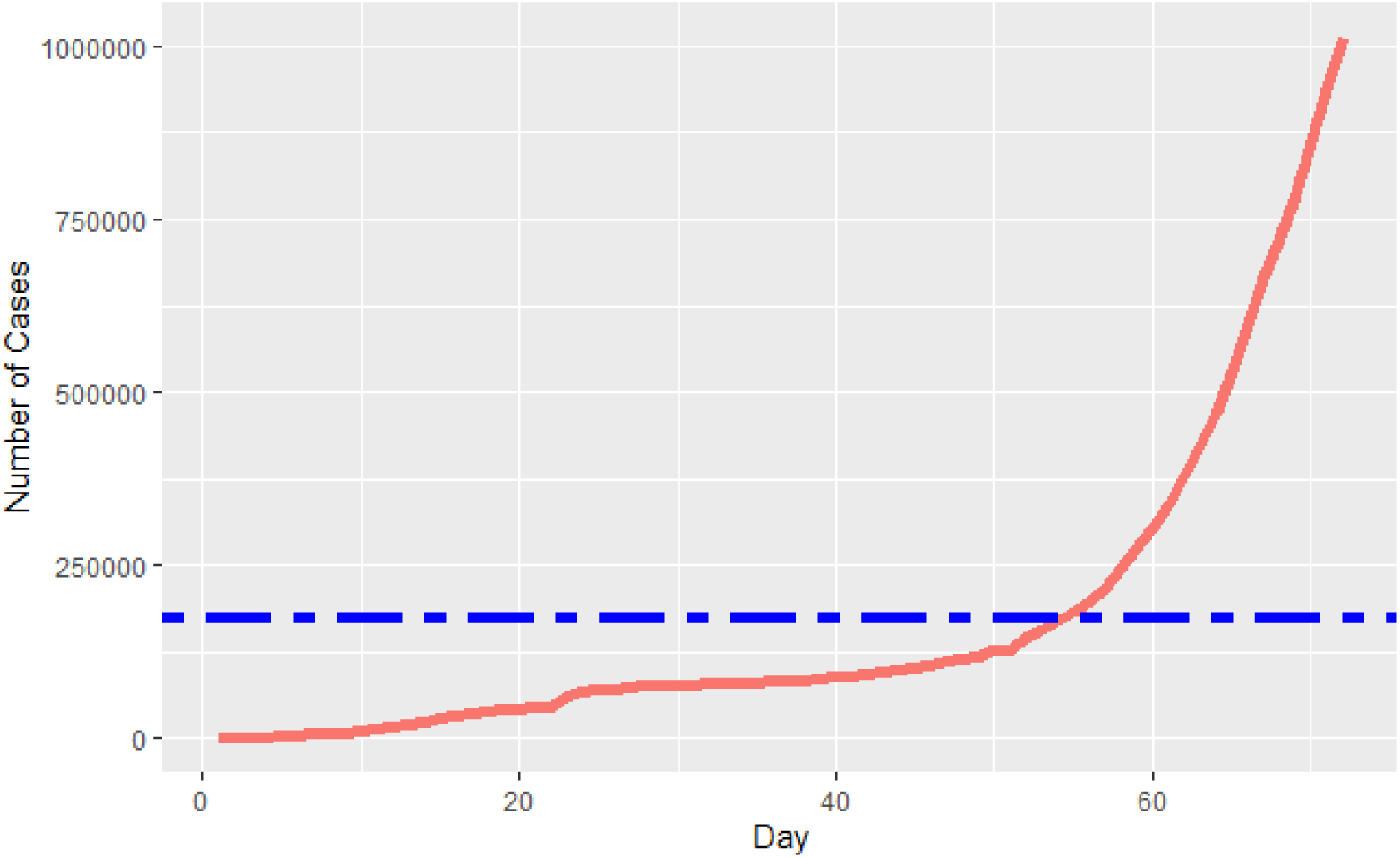
Progress of Approved Cases by Time **Data Source:** The Data Exchange (HDX)

In Figure 2, the course followed by the death cases is given by time. Similar to that in Figure 1, it was observed that there was a sharp increase at the end of the first 60 days of the epidemic in death cases according to Figure 2, and the number of death cases was above the average number of death cases.

**Figure 2:**
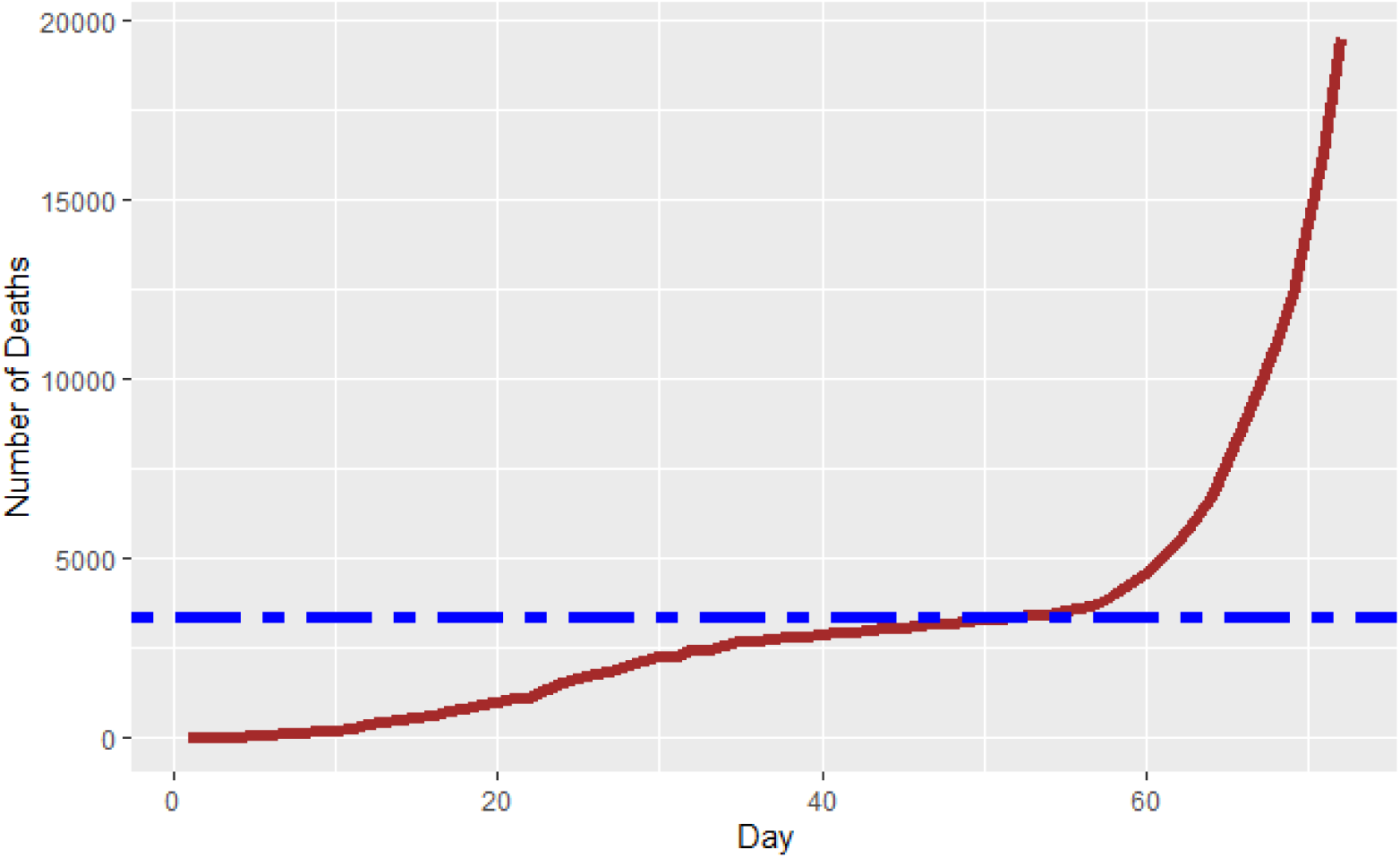
Progress of Death Cases by Time **Data Source:** The Data Exchange (HDX)

The progress of the recovered cases according to time is given in Figure 3. Unlike Figures 1 and 2, it has been observed in Figure 3 that there is a sharp increase at the end of the first 40 days of the outbreak in recovered cases, and the number of recovered cases was above the average number of recovered cases.

**Figure 3:**
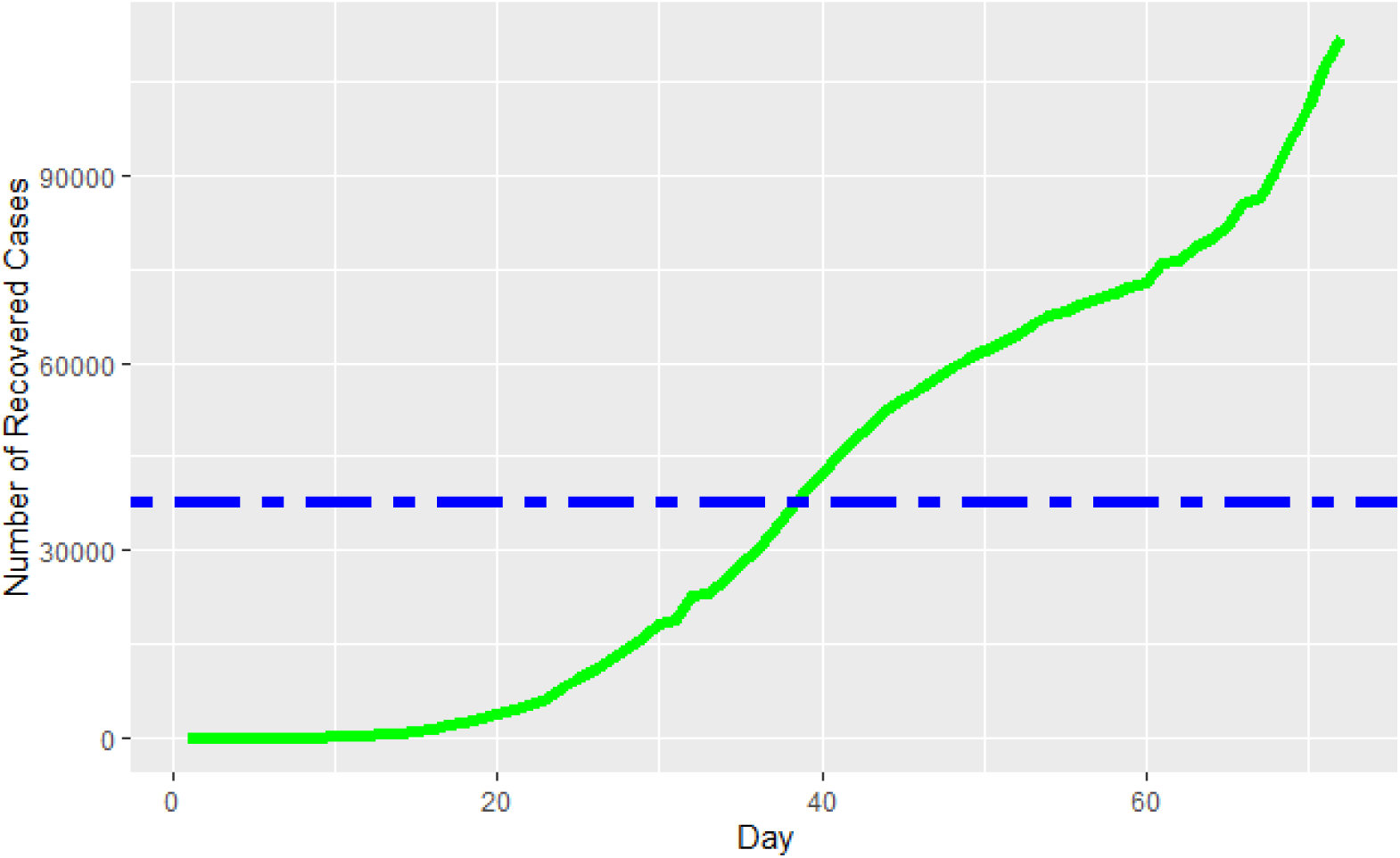
Course of Recovered Cases by Time **Data Source:** The Data Exchange (HDX)

The per-case statistics of the first 25 countries with the highest number of confirmed cases were also discussed in the timeframe from 2020-01-22 until 2020-04-22 (including this date). In this context, the number of confirmed cases and deaths per day as well as the ones of recovered cases per day was examined. The findings are given in Table 2 by country. The findings that stand out according to Table 2 are as follows:

- In the number of cases per day, USA ranked first with 3381.3 cases, followed by Italy, Spain and Germany, respectively.
- Italy ranked first in the number of deaths per day with approximately 221 cases, followed by Spain, USA, France and Iran.
- China ranked first in the number of recovered cases per day with approximately 1064.3 cases, followed by Spain, Germany, Italy and Iran.

**Table 2:** Number of Cases, Deaths and Recovered Per Day in Countries with the Highest Number of Approved Cases

| Country | Case | Death | Recovered | Day | Cases per Day | Deaths per Day | Recovered per Day |
| --- | --- | --- | --- | --- | --- | --- | --- |
| US | 243453 | 5926 | 9001 | 72 | 3381.3 | 82.3 | 125.0 |
| Italy | 115242 | 13915 | 18278 | 63 | 1829.2 | 220.9 | 290.1 |
| Spain | 112065 | 10348 | 26743 | 62 | 1807.5 | 166.9 | 431.3 |
| Germany | 84794 | 1107 | 22440 | 67 | 1265.6 | 16.5 | 334.9 |
| China | 82432 | 3322 | 76565 | 72 | 1144.9 | 46.1 | 1063.4 |
| France | 59929 | 5398 | 12548 | 70 | 856.1 | 77.1 | 179.3 |
| Iran | 50468 | 3160 | 16711 | 44 | 1147.0 | 71.8 | 379.8 |
| United Kingdom | 34173 | 2926 | 192 | 63 | 542.4 | 46.4 | 3.0 |
| Switzerland | 18827 | 536 | 4013 | 38 | 495.4 | 14.1 | 105.6 |
| Turkey | 18135 | 356 | 415 | 23 | 788.5 | 15.5 | 18.0 |
| Belgium | 15348 | 1011 | 2495 | 59 | 260.1 | 17.1 | 42.3 |
| Netherlands | 14788 | 1341 | 260 | 36 | 410.8 | 37.3 | 7.2 |
| Canada | 11284 | 138 | 0 | 68 | 165.9 | 2.0 | 0.0 |
| Austria | 11129 | 158 | 1749 | 38 | 292.9 | 4.2 | 46.0 |
| Korea. South | 9976 | 169 | 5828 | 72 | 138.6 | 2.3 | 80.9 |
| Portugal | 9034 | 209 | 68 | 32 | 282.3 | 6.5 | 2.1 |
| Brazil | 8044 | 324 | 127 | 37 | 217.4 | 8.8 | 3.4 |
| Israel | 6857 | 36 | 338 | 42 | 163.3 | 0.9 | 8.0 |
| Sweden | 5568 | 308 | 103 | 63 | 88.4 | 4.9 | 1.6 |
| Norway | 5147 | 50 | 32 | 37 | 139.1 | 1.4 | 0.9 |
| Australia | 5116 | 24 | 520 | 68 | 75.2 | 0.4 | 7.6 |
| Czechia | 3858 | 44 | 67 | 33 | 116.9 | 1.3 | 2.0 |
| Ireland | 3849 | 98 | 5 | 34 | 113.2 | 2.9 | 0.1 |
| Denmark | 3573 | 123 | 1172 | 36 | 99.3 | 3.4 | 32.6 |
| Russia | 3548 | 30 | 235 | 63 | 56.3 | 0.5 | 3.7 |

**Data Source:** The Data Exchange (HDX)

### Theoretical Framework of Models

In this section, the parameters used in model equation are firstly included. Then, case, death, recovered and net wavelength models are given respectively. The relation of wavelength equations with net wavelength equation is presented in Figure 4.

**Figure 4:**
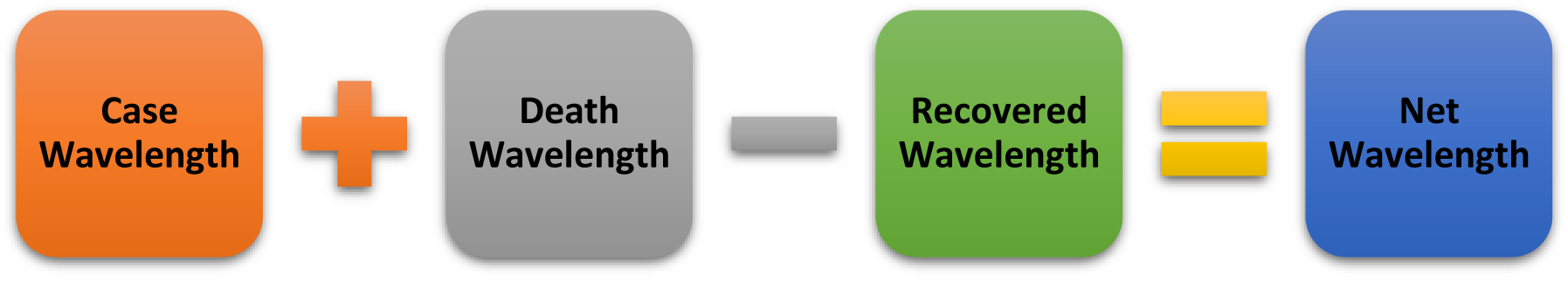
Relationship of Wavelength Equations with Net Wavelength

In order to calculate the net wavelength in equations in Figure 4, firstly, case and death wavelength equations should be calculated. Then, by calculating the recovered wavelength, net wavelength is obtained by subtracting the case and death wavelength equations. The parameters used in wavelength equations are given below.

***c***_***c***_: Approved cumulative total number of cases

***d***_***c***_: Approved cumulative total death number of cases

***t***_***c***_: Number of days since the first case was announced

***t***_***r***_: The ratio of within the total day of the year of the number of days passed since the first case announced 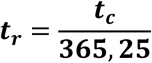

***r***_***c***_: Approved cumulative total recovered number of cases

***ln*:** Natural logarithm

***W*:** Wavelength

***W***_***c***_: Case wavelength

***W***_***d***_: Death wavelength

***W***_***r***_: Recovered wavelength

***W***_***net***_: Net wavelength

After the parameters are included, it is the recommended equations for the wavelengths. The reason for using natural logarithm (ln) in equations is that the values obtained are wanted to be normalized. The reason why the natural logarithm product coefficient is included in 1 is that ln (1) = 0.

Average is not used in wavelength equations. This is because, for example, the number of cases seen in a total of 20 days since the first epidemic case occurred in an “A” population was 2000. In another example, the number of cases seen in a total of 40 days since the first epidemic case occurred in a population “B” was 4000. In both samples, the average number of cases per day in the “A” and “B” population is equal. However, populations with different days and number of cases should be differentiated from each other. Therefore, evaluating these two populations with the same average on the same plane results in erroneous findings. These and similar situations also exist at epidemiological rates. Therefore, they are far from revealing the magnitude of the epidemic, in other words, wavelength. In general terms, as the values of the variables in the numerator and denominator grow or shrink simultaneously, there will be no differentiation. An example of this is the case-based fatality rate, which is one of the epidemiological rates. A case fatality rate (CFR) is the proportion of those who died of a particular disease at a given time. In CFR product coefficient is 100 [1]. For example, in the population “A”, the total number of cases in a given disease and at a given time is 1000, and the number of deaths is 50. In the population “B”, the total number of cases in a given disease and at a certain time is 10000, and the number of people who died is 500. In this case, the case fatality rate in these two populations is 5%.

The issue to be considered here is to reveal how many cases were reached in which time period in COVID-19 or other epidemic data sets. The task that needs to be done later is to calculate the wavelength of the outbreak using these findings. Many models developed in the study were tested in measuring the wavelength of the outbreak. Finally, it was decided to use the wavelength equations mentioned in the following sections.

### 1. Case Wavelength

Case wavelength was calculated with the help of equation (1). The higher the case wavelength (W_c_), the higher the size of the outbreak in the population.

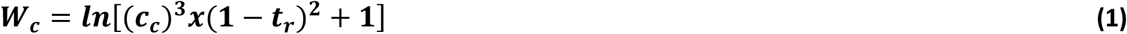

### 2. Death Wavelength

Death wavelength was calculated using equation (2). As the death wavelength (W_d_) increases, the epidemic’s killer effect increases in the population.

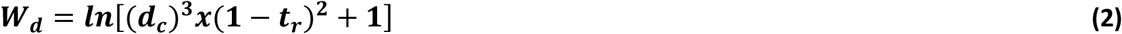

### 3. Recovered Case Wavelength

Recovered case wavelength was calculated with the help of wavelength equation (3). As recovered wavelength (W_r_) increases, the recovery rate of cases infected by the epidemic increases in the population.

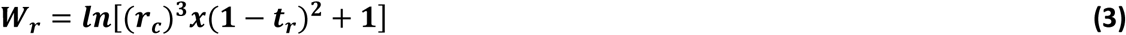

### 4. Net Wavelength

Net wavelength was calculated with the help of equation (4). Here, the net wavelength is obtained by subtracting the recovered wavelength from the sum of the case and death wavelengths. As the net wavelength increases, the net effect of the epidemic increases in the population.

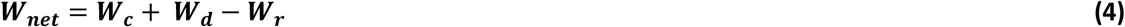

## Findings

This section includes the findings obtained from each wavelength equation mentioned in the previous sections. The findings were obtained from the COVID-19 cases, deaths and recovered cases in 181 countries during the period from 2020-01-22 until 2020-04-02 (including this date) when the first case was recorded. In the findings section, the wavelength lengths of the first 12 countries with the highest wavelength in each wavelength equation are given.

### 1. Case Wavelength (W_c_)

Findings regarding case wavelengths are presented in Table 3 comparatively by the first 12 countries with the highest wavelengths. The findings that stand out according to Table 3 are as follows:

- The USA, with the highest case wavelength, ranks first with 36.77 points, while Belgium is 12th with 28.56 points. The USA with the highest wavelength is followed by Italy, Spain and Germany, respectively.

**Table 3:** Case Wavelength (W_c_)

| Country | Rank | $c_c$ | $t_c$ | $W_c$ |
| --- | --- | --- | --- | --- |
| USA | 1 | 243453 | 72 | 36.77 |
| Italy | 2 | 115242 | 63 | 34.59 |
| Spain | 3 | 112065 | 62 | 34.51 |
| Germany | 4 | 84794 | 67 | 33.64 |
| China | 5 | 82432 | 72 | 33.52 |
| France | 6 | 59929 | 70 | 32.58 |
| Iran | 7 | 50468 | 44 | 32.23 |
| United Kingdom | 8 | 34173 | 63 | 30.94 |
| Switzerland | 9 | 18827 | 38 | 29.31 |
| Turkey | 10 | 18135 | 23 | 29.29 |
| Netherlands | 11 | 14788 | 36 | 28.60 |
| Belgium | 12 | 15348 | 59 | 28.56 |

### 2. Death Wavelength (W_d_)

Findings including death wavelengths are comparatively given by the first 12 countries with the highest wavelengths in Table 4. The findings that stand out according to Table 4 are as follows:

- Unlike case wavelength, Italy, with the highest death wavelength, ranks first with 28.24 points, while Turkey ranks 12th with 17.49 points. Italy, which has the highest death wavelength, is followed by Spain, USA and France, respectively.

**Table 4:** Death Wavelength (W_d_)

| Country | Rank | $d_c$ | $t_c$ | $W_d$ |
| --- | --- | --- | --- | --- |
| Italy | 1 | 13915 | 63 | 28.24 |
| Spain | 2 | 10348 | 62 | 27.36 |
| USA | 3 | 5926 | 72 | 25.62 |
| France | 4 | 5398 | 70 | 25.36 |
| Iran | 5 | 3160 | 44 | 23.92 |
| China | 6 | 3322 | 72 | 23.89 |
| United Kingdom | 7 | 2926 | 63 | 23.57 |
| Netherlands | 8 | 1341 | 36 | 21.40 |
| Germany | 9 | 1107 | 67 | 20.62 |
| Belgium | 10 | 1011 | 59 | 20.40 |
| Switzerland | 11 | 536 | 38 | 18.63 |
| Turkey | 12 | 356 | 23 | 17.49 |

### 3. Recovered Case Wavelength (W_r_)

Findings including recovered case wavelengths are presented by the first 12 countries with the highest wavelengths in Table 5. The findings that stand out according to Table 5 are as follows:

- Unlike other wavelengths, in recovered case wavelength, China was ranked 1st with 33.30 points, while Denmark ranked 12th with 20.99 points. China having the highest recovered case wavelenght, is followed by Spain, Germany and Italy, respectively.

**Table 5:**
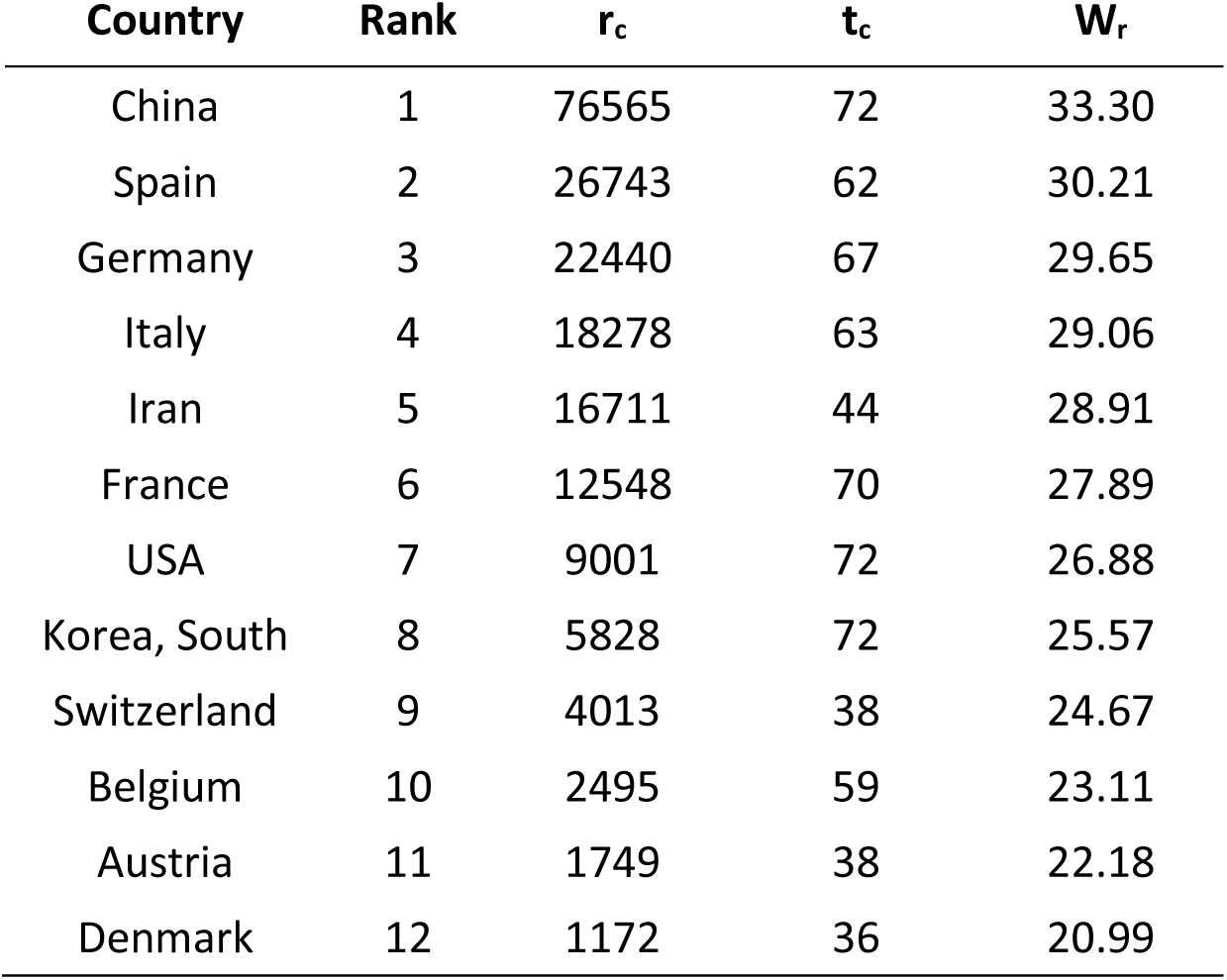
Recovered Case Wavelength (W_r_)

### 4. Net Wavelength (W_net_)

Findings related to net wavelength are presented from large to small by the first twelve countries in Table 6.Here, case and death wavelengths are summed first for the net wavelength. Then, the net wavelength was calculated by subtracting the recovered wavelength from this total. The net wavelength findings that stand out according to Table 6 are as follows:

- Unlike all other wavelengths, in net wavelength, Canada was ranked 1st with 41.95 points, while Turkey ranked 12th with 28.83 points. Canada, with the highest net wavelenght, is followed by United Kingdom, USA and Italy, respectively.

**Table 6:** Net Wavelength (W_net_)

| Country | Rank | $W_{net}$ | $W_c$ | $W_d$ | $W_r$ |
| --- | --- | --- | --- | --- | --- |
| Canada | 1 | 41.95 | 27.58 | 14.37 | 0.00 |
| United Kingdom | 2 | 39.11 | 30.94 | 23.57 | 15.39 |
| USA | 3 | 35.51 | 36.77 | 25.62 | 26.88 |
| Italy | 4 | 33.77 | 34.59 | 28.24 | 29.06 |
| Netherlands | 5 | 33.52 | 28.60 | 21.40 | 16.47 |
| Ireland | 6 | 33.49 | 24.57 | 13.56 | 4.64 |
| Spain | 7 | 31.66 | 34.51 | 27.36 | 30.21 |
| Serbia | 8 | 31.18 | 21.04 | 10.14 | 0.00 |
| Portugal | 9 | 30.51 | 27.14 | 15.84 | 12.48 |
| France | 10 | 30.05 | 32.58 | 25.36 | 27.89 |
| Brazil | 11 | 29.57 | 26.76 | 17.13 | 14.32 |
| Turkey | 12 | 28.83 | 29.29 | 17.49 | 17.95 |

## Conclusion

Within the scope of this study, mathematical models were developed to measure the extent of outbreaks in other outbreaks, especially COVID-19 pandemics, which affect countries in social, economic and many other aspects. In this way, by calculating the wavelength of the outbreaks, it is aimed to facilitate the follow-up of the outbreaks in the country and at the international level as reliable, valid and at the same time as easy as possible. At the same time, the findings from the models are expected to contribute to policy making for decision makers. On the other hand, although the developed models are designed to reveal the extent of outbreaks, they can be used in other diseases with and without infection origin.

The developed wavelength models were tested on datasets with COVID-19 cases, deaths and recovered cases in 181 countries. According to the findings obtained from the implementation, the first four countries with the highest case wavelength are USA, Italy, Spain and Germany, respectively. However, Italy ranks first in the death wavelength, followed by Spain, USA and France. On the other hand, China has taken the first place in the recovered wavelength. This country was followed by Spain and Germany and Italy, respectively. Based on all these wavelength models mentioned, net wavelength lengths are calculated. According to the net wavelengths obtained, Canada ranked first, followed by United Kingdom, USA and Italy, respectively.

Since there were no other variables other than the number of confirmed cases, deaths and recovered cases in the existing data sets, the models developed were limited to these variables. Of course, other variables can also be included in the wavelength equations mentioned as parameters, and wavelengths can be calculated.

## Data Availability

All of the data presented in this study is available on The Humanitarian Data Exchange (HDX) (https://data.humdata.org/dataset/novel-coronavirus-2019-ncov-cases).

